# The CMSES-4: A Short Scale for Pediatric Medication Self-Efficacy

**DOI:** 10.1101/2025.01.23.25320754

**Authors:** Ziying Yu, Jin Yu, Yongqiang Gao, Ting Wang, Pan Liu, Xinyu Wang, Qinghui Meng, Yibo Wu

## Abstract

This study developed and validated the Children’s Medication Self-Efficacy Scale (CMSES), a psychometric tool grounded in Bandura’s self-efficacy theory, to assess medication self-efficacy in pediatric populations. Using data from 2,258 Chinese children, the four-item scale demonstrated robust psychometric properties through exploratory factor analysis (EFA) and confirmatory factor analysis (CFA), revealing a two-factor structure (magnitude and strength) that accounted for 76% of the total variance. The scale exhibited excellent internal consistency (Cronbach’s α = 0.92). Latent profile analysis (LPA) identified three distinct self-efficacy profiles and Receiver operating characteristic (ROC) curve analysis established a cut-off score to effectively distinguish low self-efficacy cases. These findings confirm the CMSES as a reliable and valid tool for evaluating medication self-efficacy in children, offering clinicians a practical instrument to design tailored interventions aimed at improving medication adherence in pediatric care.

## 1 Introduction

Medication adherence is essential for effective treatment outcomes. Despite its critical role in clinical care, adherence to long-term medication remains poor worldwide (Foley et al., 2021; Stewart et al., 2023), which compromises treatment efficacy and increases patient mortality. This problem is particularly serious in pediatric populations, where medication use often depends on caregiver supervision and children may show lower tolerance for medication-taking due to factors such as unpleasant taste and difficulties with swallowing (Matsui, 2007). Consequently, greater attention is needed to improve medication adherence among children.

A primary predictor of medication adherence is medication self-efficacy, defined as the patient’s confidence in taking medication correctly. Research has found a correlation between medication self-efficacy and adherence across a range of chronic conditions (Walker et al., 2014; Criswell et al., 2010; Barclay et al., 2007; Molassiotis et al., 2002; Wolf et al., 2007). This association is also evident in pediatric populations, where children with higher medication self-efficacy tend to adhere more effectively to their prescribed regimens (Littlefield et al., 1992; Ott et al., 2000).

Given its central role in adherence, accurate measurement of medication self-efficacy is essential for identifying children at risk of poor adherence and for informing targeted interventions. Several medication self-efficacy scales have been developed for adult populations, including the Medication Understanding and Use Self-Efficacy Scale (Cameron et al., 2010), the Self-Efficacy for Appropriate Medication Use Scale (Risser et al., 2007), and condition-specific instruments for hypertension, diabetes, and HIV (Fernandez et al., 2008; Sleath et al., 2016; Erlen et al., 2010). However, directly applying scales developed for adults to children may introduce bias, as these scales assume cognitive and contextual capacities that may exceed the developmental level of younger children. Accordingly, there is a need for a pediatric-specific instrument to assess medication self-efficacy.

A critical step in developing such an instrument is to identify the core components of medication self-efficacy. However, the majority of existing scales lack a theoretically grounded conceptualization of this construct. In the present study, we conceptualize medication self-efficacy within Bandura’s (1977) social cognitive theory, which defines self-efficacy as individuals’ beliefs in their capability to successfully perform the actions required to achieve a desired outcome. Within this framework, self-efficacy is commonly described along three dimensions: magnitude, generality, and strength. Magnitude refers to perceived capability across varying levels of task difficulty, whereas strength measures the effort one is willing to exert when faced with disconfirming experiences. The generality dimension reflects whether confidence is limited to specific situations or extends across domains; however, it was not included in the present study, as the scale focuses exclusively on medication-related behaviors.

While both magnitude and strength are essential components of medication self-efficacy, previous scales have typically addressed only one of these. For example, the Medication Understanding and Use Self-Efficacy (MUSE) scale distinguishes between taking medication and learning about medication, primarily reflecting variations in task magnitude. In contrast, the Self-Efficacy for Appropriate Medication Use Scale (SEAMS) focuses on the strength dimension by assessing individuals’ confidence in maintaining adherence under changing life circumstances (e.g., being away from home). As these instruments assess only one dimension at a time or are confined to specific scenarios, they may be sensitive to situational changes, raising concerns about their psychometric robustness (Lamarche et al., 2018). Although one qualitative study considered both magnitude and strength dimensions simultaneously (De Geest et al., 1994), the resulting item pool was lengthy, and its psychometric validity was not formally established. Even after simplification, the scale retained 27 items (Denhaerynck et al., 2003), making it less practical in pediatric populations. To date, no validated measure has integrated both magnitude and strength dimensions of medication self-efficacy in a brief, child-appropriate format.

To address this gap, the present study aimed to develop a brief, theory-driven, and developmentally appropriate measure of medication self-efficacy for children that can be applied in both clinical screening and large-scale research settings. We developed a four-item scale based on existing instruments and expert review, designed to align with children’s cognitive development while capturing both magnitude and strength dimensions of medication self-efficacy. The scale’s reliability and validity were evaluated, and latent profile analysis (LPA) was subsequently used to examine self-efficacy distributions among Chinese children and to establish a cutoff score for identifying low medication self-efficacy. This scale provides a practical and theoretically grounded tool for identifying children at risk of poor adherence and for guiding targeted interventions in pediatric care.

## 2 Methods

### 2.1 Study design

The present research, designed as a methodological study, aimed to develop the Children’s Medication Self-Efficacy Scale and provide clinicians with a practical tool to better assess medication adherence and the effectiveness of interventions in pediatric populations.

This study consisted of three different phases: Phase1(Scale development), Phase 2(the validation process), Phase 3(cut-off point) (Fig. 1)

### 2.2 Sample

The data for this study were obtained from the cross-sectional dataset of the "Psychological and Behavioral Investigation of Chinese Residents (PBICR) 2023," a multicenter, large-sample, nationwide cross-sectional study (Zhang et al., 2025). The study was conducted from October to December 2022 in children’s hospitals throughout 22 provinces, five autonomous regions, and four municipalities in China. Investigators designated survey locations within their separate pediatric hospitals, enlisting participants aged 8 to 18 years. The identities of the participants were confirmed to confirm compliance with the inclusion criteria and absence of the exclusion criteria. If in-person surveys were viable in the hospital, researchers administered electronic questionnaires individually, accessible via a QR code scan. If in-person conditions were not satisfied, questionnaires were disseminated through communication software, and in-person surveys were done via video conversations.

Of the 3,532 subjects, 2,745 persisted following logical screening, with 2,258 finally incorporated. This sample size satisfies the criteria for stable scale evaluation as outlined by scholar DeVellis(DeVillis, 1991).The criteria for inclusion in this study were as follows: (a) patients aged 8 to 17 years; (b) children and their guardians who voluntarily participated and provided signed informed consent (guardians signed on behalf of the children); (c) comprehension of the significance of each item in the questionnaire; (d) capability to complete the questionnaire independently or with guardian assistance. Patients were excluded if they fulfilled the following criteria: (a) individuals with impaired consciousness or psychological disorders; (b) individuals with cognitive impairments.

### 2.3 Phase 1: Scale development

#### 2.3.1 Generating an item pool

This research was informed by Bandura’s self-efficacy theory. Bandura delineated three characteristics of self-efficacy: magnitude, strength, and generality. Our research utilized the first two dimensions (magnitude and strength) as the foundational framework for scale development. Through systematic literature searches using the strategy ("Self-efficacy scale" OR "self-efficacy measure" OR "self-efficacy instrument" OR "self-efficacy questionnaire") AND ("pediatric medication" OR "child medication" OR "pediatric drugs" OR "pediatric pharmacy") AND ("development" OR "construction" OR "creation" OR "validation" OR "psychometric properties" OR "reliability" OR "validity") AND ("Bandura’s self-efficacy theory" OR "Bandura self-efficacy" OR "Albert Bandura" OR "social cognitive theory" OR "social learning theory"), we comprehensively reviewed relevant studies from PubMed, Embase, ScienceDirect, and China National Knowledge Infrastructure (CNKI), through critical analysis of existing literature and reference to established medication self-efficacy scales (e.g., Self-Efficacy for Appropriate Medication Use Scale (SEAMS) (Risser et al., 2007), Medication Understanding and Use Self-Efficacy Scale (MUSE)(Cameron et al., 2010), our research team independently developed six initial items aligned with the theoretical dimensions.(Table 1)

**Table 1.** Original CMSES scale items.

| Item number | Item |
| --- | --- |
| **1** | **I find it easy to take medication daily on time.** |
| **2** | **I easily remember the correct methods of medication use** |
| 3 | I clearly understand the side effects and precautions of my medications |
| **4** | **I can still take medication on time when methods change.** |
| **5** | **Even with a busy schedule,I take medication on time.** |
| 6 | I can still take my medication on time when I'm feeling unwell |

Bolded items are retained in final CMSES scale. Item 1 and 2 are magnitude dimension , Item 4 and 5 are strength dimension

#### 2.3.2 Expert review and language adequacy

After the creation of the item pool, expert opinions were sought to assess the content validity. The experts team was composed of three paediatric nursing specialists, one paediatric psychology specialist, and one public health management specialist. The content validity of the scale was evaluated using the “Davis Technique”, a widely recognized method for evaluating content validity, was used to assess each item’s relevance and clarity. Specialists assigned scores ranging from 1 to 4 points (1=Not applicable;2=The item needs appropriate replacement;3=Suitable but requires minor modification;4=Very suitable) to determine whether the items effectively measured the relevant concept and if they should be retained in the scale. Items with a content validity Index (CVI) below 0.80 were excluded(Polit & Beck, 2006). In line with experts opinions, items 3 and 6 were removed from the scale due to a low content validity index. Then two graduate students of Chinese language performed further linguistic refining to improve semantic clarity and grammatical precision. The final scale comprised four items: two evaluating the magnitude dimension and two measuring strength. Employing a 5-point Likert scale (1 = "Strongly Disagree" to 5 = "Strongly Agree"), elevated scores signify enhanced pediatric medication self-efficacy. (Table 1)

#### 2.3.3 Pilot study

In the pilot study, the 4-item draft was conducted with 15 children undergoing treatment who met the sampling criteria but were not included in the main sample. The pilot study aimed to evaluate the readability, comprehensibility, and response time. Furthermore, the children provided positive feedback and efficiently completed the form, with an average response time of 4 min. Results from the pilot test indicated that no issues with the readability or comprehensibility of the items.

### 2.4 Phase 2: the validation process

#### 2.4.1 Validity test of the scale

For item analysis of the 4 items, the Item-total correlation (ITC) was evaluated. Following item analysis, construct validity and criterion validity were evaluated to verify the validity of the preliminary scale in this phase. EFA, CFA and convergent and discriminant validity were performed to verify the construct validity(Boateng et al., 2018).

##### 2.4.1.1 Construct validity

###### Exploratory factor analysis

Exploratory factor analysis (EFA) was performed to ascertain the scale’s structure utilizing IBM SPSS 27.0 software. Prior to executing the factor analysis, the Kaiser-Meyer-Olkin (KMO) test and Bartlett’s test of sphericity were administered. A KMO score exceeding 0.70 and a p-value below 0.05 from Bartlett’s test of sphericity signify that the data are appropriate for factor analysis.

Research demonstrates that for substantial sample sizes and non-normally distributed data, Generalized Least Squares (GLS) is the optimal approach for doing EFA (Özvarış et al., 2022). Orthogonal rotation presupposes the absence of association among components; however, this is seldom true for study variables. Consequently, methodologists advocate for the utilization of oblique rotation(Norris & Lecavalier, 2010). Given the non-normal distribution of the 2258 data points in this study and the presence of correlations among the factors, the Generalized Least Squares (GLS) method was eventually selected for factor extraction, accompanied by the optimal oblique rotation technique. Items exhibiting factor loadings exceeding 0.4 were preserved.

###### Confirmatory factor analysis

Confirmatory factor analysis (CFA) was used to verify the scale’s structure. This research employed Mplus 8.0 utilizing the greatest likelihood method to conduct confirmatory factor analysis on the sample. The model’s fit was assessed using five indices: χ²/df, CFI, TLI, RMSEA, and SRMR. The standards for an acceptable model fit were χ²/df<3, CFI>0.9, TLI>0.9, RMSEA<0.08, and SRMR<0.08. Meeting these requirements signifies an adequate model match.

###### Convergent and Discriminant Validity

Convergent validity refers to the degree of concordance between observed variables that measure latent factors. The convergent validity of the scale was assessed by the Average Variance Extracted (AVE) and Composite Reliability (CR) metrics, convergent validity is considered good when AVE>0.5 and CR>0.7.

Discriminant validity refers to the degree to which the difference between independent latent variables is indicated(Uhm, 2023). To evaluate discriminant validity, we computed the correlations across latent variables and contrasted them with the square root of the Average Variance Extracted (AVE). Discriminant validity is established when all correlation coefficients are inferior to their respective √AVE, signifying that each factor is separate from the others.

##### 2.4.1.2 Criterion validity

Criterion validity assesses the association between the present scale and a recognized scale, where higher correlation coefficients signify enhanced criterion validity and increased reliability of the scale(Yi et al., 2006). Criterion validity includes both concurrent validity and predictive validity, primarily differing in the time aspect.

This study employed the Short General Self-Efficacy Scale (SGSES) to assess concurrent validity. The SGSES was selected due to its brevity and dependability, demonstrating strong validity and reliability, hence offering significant convenience for researchers and participants alike. Furthermore, it exhibits adaptability and relevance within the Chinese demographic. The SGSES comprises three items, creating a unidimensional scale, with scores ranging from 0 to 15; elevated values signify greater self-efficacy. The Cronbach’s α coefficient of the SGSES is 0.876, and in this study, the Cronbach’s α coefficient was found to be 0.956.

#### 2.4.2 Reliability test of the scale

The reliability of the scale was evaluated by calculating the internal consistency of the items. Internal consistency denotes the reliability and uniformity of scores derived from the scale during evaluation(Yuxi Liu et al., 2023). We assessed the internal consistency of the scale utilizing Cronbach’s α coefficient. A Cronbach’s α coefficient beyond 0.8 signifies outstanding reliability; above 0.7 is deemed acceptable; above 0.6 implies the scale need revision although retains some utility; a coefficient below 0.6 shows the scale necessitates redesign (Yew et al., 2023).

We evaluated the split-half reliability of the scale. This entailed partitioning the scale into two segments, achievable either by bisecting it into the first and second halves or by segregating it between odd and even pieces. The Pearson correlation coefficient between the scores of the two sections was subsequently computed. A split-half reliability coefficient of 0.8 is typically necessary.

### 2.5 Phase 3: Cut-off point

#### 2.5.1 Latent profile analysis

In contrast to variable-centered statistical methods that often categorize based on averages and proportions, Latent Profile Analysis (LPA) adopts a person-centered approach. In the lack of a definitive "gold standard" for measurement instruments, LPA is frequently employed to determine cutoff values for scales(Wu et al., 2023). It utilizes the Latent Profile Model (LPM) to discern latent subgroups from a collection of continuous variables and categorize individuals into many different profiles.

This research employed Mplus 8.0 software to perform Latent Profile Analysis on four items from the Children’s Medication Self-Efficacy Scale. Commencing with a singular profile, models were progressively augmented, calibrated, and evaluated according to metrics including the Akaike Information Criterion (AIC), Bayesian Information Criterion (BIC), Sample-Size Adjusted BIC (aBIC), Bootstrap Likelihood Ratio Test (BLRT), Lo–Mendell–Rubin Test (LMRT), and entropy, until the optimal model was discerned(Tein et al., 2013). Lower values of AIC, BIC, and aBIC signify a superior fit, although these metrics often diminish as the number of profiles increases. An entropy value of ≥0.80 signifies superior classification quality(Li et al., 2020). The BLRT and LMRT are employed to assess disparities between the two models (k-profile versus k-1 profile), with P < 0.05 indicating the superiority of the k-model(Nylund et al., 2007).

#### 2.5.2 Receiver operating characteristic analysis

ROC analysis was performed utilizing SPSS 27.0. The classification performance was assessed using measures like the Area Under the ROC Curve (AUC), sensitivity, specificity, and Youden’s index. AUC values approaching 1 signify enhanced diagnostic efficacy. The ideal cutoff value can be established using Youden’s index.

### 2.6 Ethical considerations

PBICR 2023 complied with the World Medical Association’s Declaration of Helsinki, obtained ethical approval from Beijing Children’s Hospital Capital Medical University (approval no: ChiCTR2300070667). All participants in PBICR submitted written informed consent before to engaging in the survey.

## 3 Results

### 3.1 Socio-demographic characteristics

A total of 3,532 questionnaires were disseminated, with 2,745 retained following logical screening, resulting in a response percentage of 77.72%. In conclusion, 2,258 questions were incorporated into the final analysis. Among the respondents, 48.3% were male; the predominant age group was 13-17 years (74.4%); a comparatively higher percentage had agricultural household registrations (52.7%); the largest cohort consisted of primary school students (39.9%), whereas the smallest cohort comprised undergraduate students (1.5%) (see Table 2).

**Table 2.** General Characteristics of Participants(N=2258)

| Variables | Mean ± SD or n (%) |
| --- | --- |
| Age (years) | 14.04±2.49 |
| Gender |  |
| Male | 1091 (48.3) |
| Female | 1167 (51.7) |
| Household registration type |  |
| Non-agricultural | 1068 (47.3) |
| Agriculture | 1190 (52.7) |
| Education level |  |
| Primary school | 902 (39.9) |
| Secondary school | 585 (25.9) |
| High school degree | 236 (10.5) |
| Technical secondary school | 463 (20.5) |
| education |  |
| Associate degree | 38 (1.7) |
| Bachelor degree | 34 (1.5) |

### 3.2 Item analysis

The correlation coefficients between the scale items and total scale were found to range from 0.909 to 0.940(p<.001).

### 3.3 Results of validity analysis

#### 3.3.1 Content validity

The I-CVI of the 6 preliminary questions ranged from 0.60 to 1.00, and the S-CVI was 0.90. Among the initial questions, two with an ICVI of 0.80 or less were deleted. 4 preliminary items were finalized.

#### 3.3.2 Construct validity

##### Exploratory factor analysis

Bartlett’s test of sphericity revealed a statistically significant difference (χ²=6540.24; P<0.001), and the Kaiser-Meyer-Olkin (KMO=0.84) value exceeded 0.60 (refer to Table 3). The results of these two tests indicate that the data are appropriate for factor analysis. Consequently, we employed Generalized Least Squares (GLS) and optimal oblique rotation to extract two fixed common factors.

**Table 3.** KMO and Bartlett’s Test (N=2258)

|  |  |
| --- | --- |
| **Kaiser–Meyer–Olkin measure of sampling adequacy** | 0.844 |
| Bartlett's test of sphericity |  |
| Approx. chi-square | 6540.243 |
| df | 6 |
| Sig | 0.000 |

N=2258.df=Degrees of freedom

Items were eliminated according to the following criteria: factor loadings below 0.40 and elevated cross-loadings (loadings exceeding 0.30 on two or more factors). The analysis indicated that all four items exhibited factor loadings exceeding 0.40 without any cross-loadings, hence all items were kept.

Table 4 displays the factor loadings for the two-factor structure: Magnitude dimension (loading range of 0.47 to 0.81) and strength dimension (loading range of 0.53 to 0.81). Factor 1 comprises items 1 and 2, which denote the magnitude dimension of medication self-efficacy, and Factor 2 has items 3 and 4, symbolizing the strength dimension. These two prevalent factors accounted for 76% of the overall variance.

**Table 4.** Rotated Factor Loadings for the scale(N=2258)

| **Items** | **Factor1** | **Factor2** |
| --- | --- | --- |
| 1.It's easy for me to take medicine on time every day | 0.472 |  |
| 2.It's easy for me to remember the correct way to take all my medications | 0.806 |  |
| 3.When my medication changed, I was able to take it on time |  | 0.425 |
| 4.When I am busy with school work, I can also take my medicine on time |  | 0.079 |

##### Confirmatory factor analysis

We performed confirmatory factor analysis with the greatest likelihood technique. Table 5 presents the five fit indices for the Confirmatory Factor Analysis (CFA): χ²=8.059, CFI=0.999, TLI=0.995, RMSEA=0.056, and SRMR=0.003. Aside from the comparatively elevated χ² value, all other indices exhibited a satisfactory match. Nonetheless, this is not a significant issue as a substantial sample size might exaggerate χ² and produce in misleading positive outcome(Yuping Liu et al., 2023).

**Table 5.**
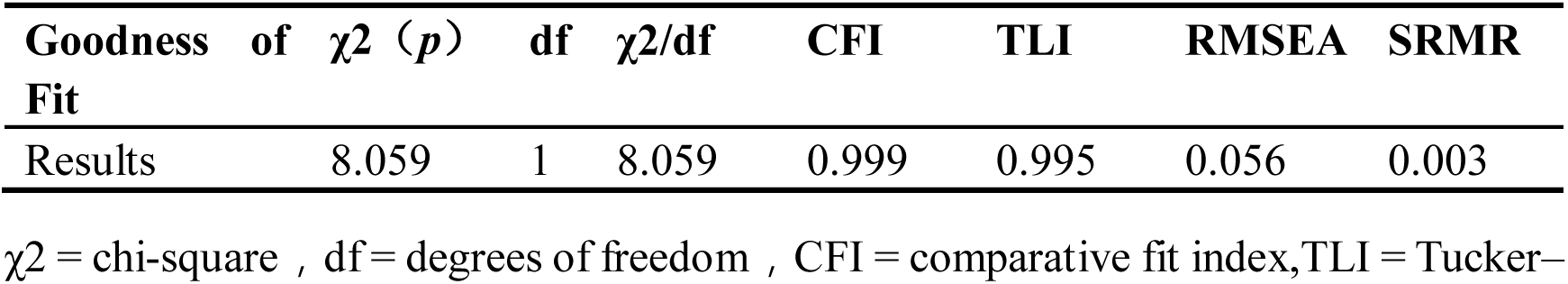
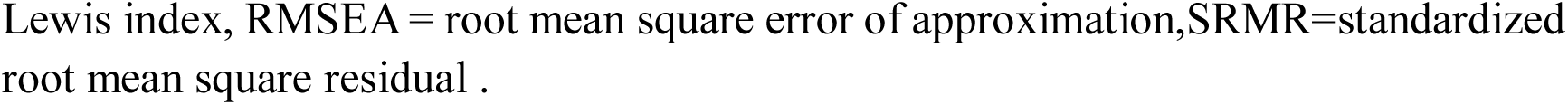
Model fit of the scale(N=2258)

| **Goodness of Fit** | **$\chi^2$ (p)** | **df** | **$\chi^2/df$** | **CFI** | **TLI** | **RMSEA** | **SRMR** |
| --- | --- | --- | --- | --- | --- | --- | --- |
| Results | 8.059 | 1 | 8.059 | 0.999 | 0.995 | 0.056 | 0.003 |

$\chi^2$ = chi-square , df = degrees of freedom , CFI = comparative fit index,TLI = Tucker–
Lewis index, RMSEA = root mean square error of approximation, SRMR = standardized root mean square residual .

##### Convergent and Discriminative Validity

The AVE values for the two factors exceeded 0.50 (magnitude dimension=0.77; strength dimension=0.85), and the CR values surpassed 0.70 (magnitude dimension=0.93; strength dimension=0.95), signifying robust convergent validity. Furthermore, the √AVE for each element exceeded the absolute values of the correlation coefficients among the factors, indicating that internal correlations surpassed external correlations. This signifies that the measure possesses strong discriminant validity (Table 6).

**Table 6.** Convergent and discriminative validity(N=2258)

|  | **AVE** | **CR** | **Factor 1** | **Factor 2** |
| --- | --- | --- | --- | --- |
| Factor 1 | 0.774 | 0.928 | 0.870<sup>a</sup> |  |
| Factor 2 | 0.847 | 0.955 | 0.832<sup>b</sup> | 0.920<sup>a</sup> |

Factor1 =magnitude dimension , Factor2 =strength dimension
AVE, average variance extracted; CR, composite reliability.
a Square root of AVE.
b Absolute value of the correlation coefficient between factors.

#### 3.3.3 Criterion validity

A negative association exists between SGSES and the Medication Self-Efficacy Scale for Children (r=-0.13, P<0.001). Every dimension exhibited a negative association with the overall score of SGSES.

### 3.4 Results of reliability analysis

The aggregate Cronbach’s α coefficient for the scale was 0.92, with the magnitude dimension at 0.83 and the strength dimension at 0.88. This signifies that the scale possesses exceptional reliability and outstanding internal consistency. The split-half reliability showed that Cronbach’s α for the first half of the scale was 0.83 and for the second half was 0.88, both greater than 0.8, demonstrating good split-half reliability.

### 3.5 Results of Latent profile analysis

We have developed potential models utilizing Latent Profile Analysis (LPA), and the fit indices for the five models are displayed in Table 7. AIC, BIC, and aBIC diminish with the rise in the number of profiles. All entropy values exceed 0.90. With the exception of the LMR p-value in the 5-Profile model, which exceeds 0.05, the p-values for the LMR and BLRT in the other profiles are statistically significant. After evaluating model fit and the practical context of this study, the 3-Profile model is determined to be the most suitable for the current sample (Table 8).

**Table 7.** Model fit indices for one- to five-latent prrofile solutions.

| **Class number** | **AIC** | **BIC** | **aBIC** | **pLMR**<br>**p-value** | **pBLRT**<br>**p-value** | **Entropy** |
| --- | --- | --- | --- | --- | --- | --- |
| 1 | 27675.206 | 27675.206 | 27695.566 |  |  |  |
| 2 | 22627.754 | 22702.143 | 22660.84 | <0.001 | <0.001 | 0.925 |
| 3 | 20605.858 | 20708.858 | 20651.669 | <0.001 | <0.001 | 0.928 |
| 4 | 18608.177 | 18739.789 | 18666.714 | 0.0013 | <0.001 | 0.959 |
| 5 | 17826.67 | 17986.892 | 17897.932 | 0.3071 | <0.001 | 0.996 |

**Table 8.** Average latent profile probabilities for most likely latent profile membership (row) by latent profile (column).

| **Latent class** | **Latent class membership** |  |  |
| --- | --- | --- | --- |
|  | **1 ( 151 )** | **2 ( 931 )** | **3 ( 1176 )** |
| 1 | 0.952 | 0.010 | 0.039 |
| 2 | 0.026 | 0.974 | 0.000 |
| 3 | 0.020 | 0.000 | 0.980 |

The columns refer to latent profile, and the rows refer to most likely profile membership. Profile 1 = low medication self-efficacy; Profile 2 = moderate medication self-efficacy; Profile 3 = high medication self-efficacy.

Figure 2 illustrates the distribution of scale components inside the 3-Profile model. Class 1, constituting 6.9% of the sample (n=151), is designated as the "Low Self-Efficacy" group. Class 2, comprising 40.5% of the sample (n=931), is designated as the "Moderate Self-Efficacy" group. Class 3, comprising 52.6% of the sample (n=1176), is designated as the "High Self-Efficacy" group. The average latent class probabilities for the most probable latent class membership (0.952, 0.974, and 0.980, respectively) suggest that the model’s classification is valid and demonstrates strong discriminative capability.

### 3.6 Results of Receiver operating characteristic analysis

According to the findings of the LPA, those categorized in the "Low Self-Efficacy" group are designated as "cases," whilst those in the "Moderate Self-Efficacy" and "High Self-Efficacy" categories are labeled as "non-cases"(Fu et al., 2022). We generated the ROC curve for the Medication Self-Efficacy Scale for Children utilizing this binary classification. The AUC value is 99.90%, signifying exceptional predictive capacity for children’s self-efficacy (Figure 3). Table 8 displays the diagnostic signs for the latent cutoff values, identifying the ideal cutoff value for the Medication Self-Efficacy Scale for Children as 16, which aligns with the highest Youden index (0.979), a sensitivity of 0.979, and a specificity of 1(Table 9).

**Table 9.** Criterion values and coordinates of ROC Curve for medication self-efficacy.

| **Criterion** | **Sensitivity** | **Specificity** | **Youden's index** |
| --- | --- | --- | --- |
| 12.5 | 1 | 0.733 | 0.733 |
| 13.5 | 1 | 0.808 | 0.808 |
| 14.5 | 1 | 0.913 | 0.913 |
| **15.5\*** | **0.979\*** | **1\*** | **0.979\*** |
| 16.5 | 0.642 | 1 | 0.642 |
| 17.5 | 0.574 | 1 | 0.574 |
| 18.5 | 0.503 | 1 | 0.503 |

## 4 Discussion

This study developed and validated a brief and efficient medication self-efficacy scale specifically designed for children. The content of the scale is based on Bandura’s self-efficacy theory and includes four items that cover the magnitude and strength dimensions of self-efficacy. This allows for the quick screening and assessment of children’s confidence in taking medication and using it correctly in various situations. The scale demonstrated good reliability and validity, making it useful for predicting children’s medication adherence and evaluating the effectiveness of medication interventions in pediatric patients with chronic illnesses.

Unlike previous studies on the development of medication self-efficacy scales, this research is grounded in a clear theoretical framework and provides a well-defined conceptualization of medication self-efficacy. We propose that medication self-efficacy should encompass the magnitude and strength dimensions of self-efficacy, i.e., the child’s confidence in handling medication tasks of varying difficulty under different life circumstances. We selected two representative items for each dimension based on expert recommendations: for the magnitude dimension, we included tasks of varying difficulty, specifically "taking medication on time" and "remembering all correct methods of medication use." For the strength dimension, we aimed to measure the patient’s perceived effort in adhering to medication when capacity is limited, choosing the items "changes in medication methods" and "busy lifestyle." By selecting these four items, we aim to comprehensively cover the two dimensions of medication self-efficacy, thereby achieving a more accurate and stable measurement of medication self-efficacy. This, in turn, enhances the predictive power of this construct in relation to medication adherence.

To assess the psychometric features of the scale in pediatric patients, we performed an internal consistency reliability analysis, indicating that the overall scale (Cronbach’s α=0.92) and both subscales (Cronbach’s α>0.83) had excellent Cronbach’s alpha coefficients. This signifies that the items within the scale reliably assess the intended constructions.

For the validation of construct validity, during the development of the scale, we aimed for the four items to represent the two dimensions of self-efficacy. Factor analysis revealed that items 1 and 2, as well as items 3 and 4, were explained by two distinct factors, aligning with our expectations. The Average Variance Extracted (AVE) values showed that the internal correlations of these two factors were greater than their external correlations, indicating good discriminant validity among the different items of the scale. We believe that these two factors represent the magnitude and strength dimensions of medication self-efficacy, respectively.

This scale provides a comprehensive measurement of the different dimensions of medication self-efficacy, which allows for a more comprehensive understanding of the framework of medication self-efficacy. The differentiation between these dimensions is essential, as starting a medication regimen does not necessarily mean that the child can take the correct dosage(Menditto et al., 2021). In this scale, the magnitude dimension represents the difficulty level of the medication task, where correctly remembering all medication instructions is more challenging than merely taking the medication on time. As the number of medications increases, the complexity of the task also increases, potentially leading to a decrease in medication adherence(Knight et al., 2001). Using two items helps capture the child’s performance across both simple and difficult medication tasks. The strength dimension of medication adherence reflects the effort a child puts into taking their medication. Adherence becomes more challenging when daily life is busy or medication routines change (Lehane & McCarthy, 2007). In such situations, self-efficacy may decline, as children often have less energy and focus to remember their medication, reducing their confidence in taking it correctly. This scale’s measurement covers the magnitude and strength dimensions, providing a more comprehensive reflection of medication self-efficacy, which in turn leads to better stability and accuracy.

Upon investigating the correlation between general self-efficacy and medication self-efficacy in children, we unexpectedly identified a strong negative association. This finding suggests that general self-efficacy may not necessarily be effective in the context of medication adherence. One possible explanation for this negative correlation is that children with high medication self-efficacy might have more severe health conditions, which may contribute to reduced overall self-efficacy. The specific mechanisms underlying this relationship require further investigation and validation.

We performed a latent profile analysis to investigate the distribution of medication self-efficacy among Chinese pediatric patients. The results indicate that the level of medication self-efficacy among the participants can be categorized into three groups: most participants had high (52.6%) or moderate (40.5%) self-efficacy, while 6.9% had low self-efficacy. We believe that children with low medication self-efficacy are more prone to medication non-adherence; therefore, they should be the primary focus of healthcare professionals for intervention.

To establish a cutoff value to identify children with low medication self-efficacy, we performed a receiver operating characteristic (ROC) analysis. It appears that a cutoff value of 15.5 yielded the highest Youden’s index, which means optimized specificity and sensitivity in distinguishing the low self-efficacy group. Thus, the cutoff value for this scale is set at 16: participants scoring greater than or equal to 16 are considered to have potentially low medication self-efficacy and should undergo further evaluation and intervention.

This scale can be used for quick assessment of medication self-efficacy in children who need to adhere to their medication. It assists in identifying individuals with low self-efficacy who are at an increased risk of non-adherence during discharge evaluations and follow-up monitoring. These children can then be identified for interventions (da Rocha Mendes et al., 2024; Iio et al., 2017; Shih & Cohen, 2020), which may include medication education for them or their caregivers, remote medication monitoring, and further support. Consequently, we can more efficiently mitigate non-adherence, diminish medication hazards, and enhance treatment outcomes. However, our study still have some limitations. First, the scale examines general medication self-efficacy without distinguishing between specific conditions, medication duration, frequency, or quantity. Longer disease courses, higher medication quantities, and fluctuating frequencies can increase the difficulty of medication adherence potentially reducing patients’ adherence and their self-efficacy in managing their medication. Future research could validate the scale’s applicability across different pediatric conditions. Second, this scale includes representative scenarios to reflect different strength and magnitude of medication self-efficacy. However, the item descriptions are based on experience and expert recommendations, so it remains to be verified whether they effectively measure children’s medication self-efficacy levels. Third, like many studies on the development of medication self-efficacy scales, this research only measured self-efficacy at a single cross-sectional time point, without examining whether this measurement can predict long-term medication adherence. Longitudinal validation is needed to enhance the reliability of the scale.

## Data Availability

All data produced in the present study are available upon reasonable request to the authors

